# The Effect of Famotidine on Hospitalized Patients with COVID-19: a Systematic Review and Meta-analysis

**DOI:** 10.1101/2021.03.14.21253537

**Authors:** Leonard Chiu, Max Shen, Ronald Chow, Chun-Han Lo, Nicholas Chiu, Austin Chen, Hyun Joon Shin, Elizabeth Horn Prsic, Chin Hur, Benjamin Lebwohl

## Abstract

**Introduction:** Famotidine is a competitive histamine H2-receptor antagonist most commonly used for gastric acid suppression but thought to have potential efficacy in treating patients with COVID-19. The aims of this systematic review and meta-analysis are to summarize the current literature and report clinical outcomes on the use of famotidine for treatment of hospitalized patients with COVID-19.

**Methods:** Five databases were searched through February 12, 2021 to identify observational studies that reported on associations of famotidine use with outcomes in COVID-19. Meta-analysis was conducted for composite primary clinical outcome (e.g. rate of death, intubation, or intensive care unit admissions) and death separately, where either aggregate odds ratio (OR) or hazard ratio (HR) was calculated.

**Results:** Four studies, reporting on 46,435 total patients and 3,110 patients treated with famotidine, were included in this meta-analysis. There was no significant association between famotidine use and composite outcomes in patients with COVID-19: HR 0.63 (95% CI: 0.35, 1.16). Across the three studies that reported mortality separated from other endpoints, there was no association between famotidine use during hospitalization and risk of death - HR 0.67 (95% CI: 0.26, 1.73) and OR 0.79 (95% CI: 0.19, 3.34). Heterogeneity ranged from 83.69% to 88.07%.

**Conclusion:** Based on the existing observational studies, famotidine use is not associated with a reduced risk of mortality or combined outcome of mortality, intubation, and/or intensive care services in hospitalized individuals with COVID-19, though heterogeneity was high, and point estimates suggested a possible protective effect for the composite outcome that may not have been observed due to lack of power. Further RCTs may help determine the efficacy and safety of famotidine as a treatment for COVID-19 patients in various care settings of the disease.

## INTRODUCTION

Coronavirus disease 2019 (COVID-19) is predominantly a respiratory illness caused by severe acute respiratory syndrome coronavirus 2 (SARS-COV-2) that first arose in December 2019 in Wuhan, China (1, 2). After one year, COVID-19 remains an ongoing pandemic that is uncontrolled in many parts of the world. Continued optimization of medical therapy remains essential in combating COVID-19. For hospitalized patients with severe disease, current therapeutic options include dexamethasone, remdesivir, convalescent plasma, and monoclonal antibodies (Bamlanivimab, Casirivimab-Imdevimab) depending on the degree of oxygen supplementation, respiratory support, as well as the specific clinical situation (3-10). Bamlanivimab and Casirivimab-Imdevimab have also received emergency use authorization from the United States Food and Drug Administration for outpatients with mild to moderate COVID-19. Pharmacologic treatment for patients prior to hospitalization remains sparse (10).

Famotidine is a competitive histamine H2-receptor antagonist. Its main pharmacodynamic effect is the inhibition of gastric acid secretion (11). In February 2020, a study by Wu *et al* (12), used computational methods to predict structures of proteins encoded by the SARS-CoV-2 genome in order to identify available drugs that may be repurposed to treat COVID-19. Famotidine was found to be a potential candidate that may inhibit 3chymotrypsin-like protease (3CLpro), a viral enzyme necessary for SARS-CoV-2 viral replication. Subsequently, several studies have reported on the use of famotidine in treating COVID-19 patients (13-18). Specifically, Freedberg *et al* and Mather *et al* (14, 15), found that in patients hospitalized with COVID-19, famotidine use was associated with a reduced risk of clinical deterioration leading to intubation or death; however, other observational studies did not find a reduced risk of mortality, intensive care unit admission, and/or intubation with the use of famotidine in patients with COVID-19 (13, 16-18).

To our knowledge, two meta-analyses on the use of famotidine in patients with COVID-19 have been published (19, 20). However, these meta-analyses may have introduced heterogeneity in patient population due to the inclusion of studies with non-hospitalized patients diagnosed with COVID-19 (13, 18). Additionally, one study (19) did not include all existing evidence published in the literature to date (16).

We therefore conducted an updated systematic review and meta-analysis with the aims to summarize current literature on the use of famotidine for COVID-19 and report clinical outcomes in only hospitalized patients with COVID-19 treated with famotidine.

## METHODS

### Search Strategy

Five databases, namely Ovid Medline, Embase, Cochrane Central Register of Controlled Trials (CENTRAL), medRxiv, and researchsquare, were searched through to February 12, 2021 (Appendix 1).

### Study Eligibility

All identified articles from the database search underwent screening, where two reviewers (LC, RC) independently assessed articles. During level 1 screening, articles were screened by their title and abstract, and were eligible for further screening if they reported on famotidine in patients with COVID-19. Studies subsequently underwent level 2 screening where their full-texts were assessed to determine eligibility based on whether the paper reported on a clinical dataset. Articles categorized as case reports, case series, reviews, or non-clinical studies were excluded. The remaining eligible studies went through a final round of assessment for quantitative synthesis, and were included in this systematic review and meta-analysis if they reported an adjusted relative risk measure of mortality and/or a composite clinical outcome for famotidine relative to non-famotidine users in hospitalized patients only.

If disagreements occurred between the two reviewers at any stage, a discussion occurred, and consensus achieved for a final decision. If discrepancies could not be resolved, a third reviewer (MS) was consulted to help achieve consensus.

### Quantitative Synthesis

As mentioned, adjusted relative risk ratios for mortality and/or another primary composite clinical outcome were extracted from each eligible article in our review. Furthermore, we noted sample size, study design, patient population, mean/median age, percentage male, percentage famotidine users, and adjusted confounding variables.

Quantitative synthesis was also done independently by two reviewers (LC, RC), and a third reviewer was consulted to resolve discrepancies when they arose (MS).

### Risk of Bias Assessment

The Risk Of Bias In Non-randomized Studies – of Interventions (ROBINS-I) tool, developed by the Cochrane Bias Methods Group, was used to assess risk of bias for all observational studies included in this meta-analysis (21). Primary assessment was conducted by one reviewer (C-HL), and subsequently re-assessed by two reviewers (LC, MS).

### Statistical Analysis

A meta-analysis was conducted by subgroups of whether patients took famotidine prior to or during hospitalization. Within subgroups, meta-analysis was conducted based on the type of relative risk ratio reported. Odds ratios (ORs) were aggregated to generate a summary OR, and hazard ratios (HRs) were aggregated to generate a summary HR. The primary outcome that was meta-analyzed was a composite outcome of mortality, intubation, or intensive care unit admission. The secondary outcome aggregated was the mortality rate separated from other composite outcomes. A random-effects DerSimonian-Laird analysis model was used, as there was high heterogeneity. A *p*-value of less than 0.05 was deemed as the threshold for statistical significance.

Due to the limited number of studies that reported results for each outcome measure, we did not assess for publication bias with a funnel plot and Egger’s test. All analyses were conducted using Stata 16.1.

## RESULTS

106 articles were located through database search and three additional articles were located through backward reference search. After duplication removal, 76 unique articles remained and underwent level 1 screening, yielding 28 articles that underwent level 2 screening. Eight articles reported on clinical dataset and therefore were eligible for possible quantitative synthesis. However, only six articles reported adjusted relative risk ratios (13-18), and two of them were excluded as they consisted of both inpatients and outpatients diagnosed with COVID-19 (13, 18). Therefore, four studies were included in this systematic review and meta-analysis. The PRISMA flow diagram is presented in Appendix 2.

Key characteristics for included studies are presented in Table 1. All studies were retrospective cohort studies in hospitalized patients with COVID-19 (14-17). While Freedberg *et al* and Mather *el al* were single-center studies conducted in the United States, the other two were multicenter studies. Mather *et al* defined a famotidine user as one who used famotidine within 7 days prior to or after the date of hospital admission and/or COVID-19 screening, while the other three studies defined famotidine users as those who were given famotidine during their hospitalizations, most commonly within 24 hours of admission. Additionally, Freedberg *et al* and Yeramaneni *et al* excluded patients who died or were intubated within 48 hours of admission, whereas Shaoibi *et al* excluded patients who received intensive care services at or up to 30 days prior to admission.

**Table 1.** Study Characteristics

| Study | Sample Size | Study Design | Patient Population | ICU At Study Enrollment? | Mean/Median Age | % Male | Definition of Famotidine Use | % Famotidine Users | Primary Outcomes | Adjusted Covariates |
| --- | --- | --- | --- | --- | --- | --- | --- | --- | --- | --- |
| **Freedberg *et al*** | 1620 | Retrospective Cohort | COVID-19 diagnosed patients | No | NR | 44 | Famotidine during hospitalization | 5.2 | Composite of death or intubation | Age, sex, race/ethnicity, BMI, comorbidities, initial oxygen requirement |
| **Mather *et al*** | 878 | Retrospective Cohort | COVID-19 diagnosed patients | No | 67 +/- 16 | 54.7 | Famotidine before and during hospitalization | 9.5 | Composite of death or ventilation | Age, sex, race, smoking status, BMI, comorbidities, national early warning score |
| **Shoaibi *et al*** | 36779 | Retrospective Cohort | COVID-19 diagnosed patients | No | NR | NR | Famotidine during hospitalization | 4.9 | Composite of death or intensive services (ventilation, tracheostomy, or ECMO) | Age, gender, comorbidities |
| **Yeramaneni *et al*** | 7158 | Retrospective Cohort | COVID-19 diagnosed patients | No | 57.9 +/- 19.3 | 49.1 | Famotidine during hospitalization | 15.7 | 30-day all-cause mortality | Age, sex, race/ethnicity, BMI, comorbidities, WHO severity, smoking status, medications |

Legend: NR – not reported

Risk of bias assessment via ROBINS-I is presented in Figure 1. In general, there are concerns regarding residual confounding due to the observational nature of all the included studies. Given that famotidine could be used prior to hospitalization and that none of the studies adopted a new user study design, the start of follow-up and the start of exposure might not coincide for famotidine users. This was the main cause of the moderate risk of selection bias. Publication bias could not be assessed via funnel plot or Egger’s test due to the limited number of studies for each outcome measure.

**Figure 1.**
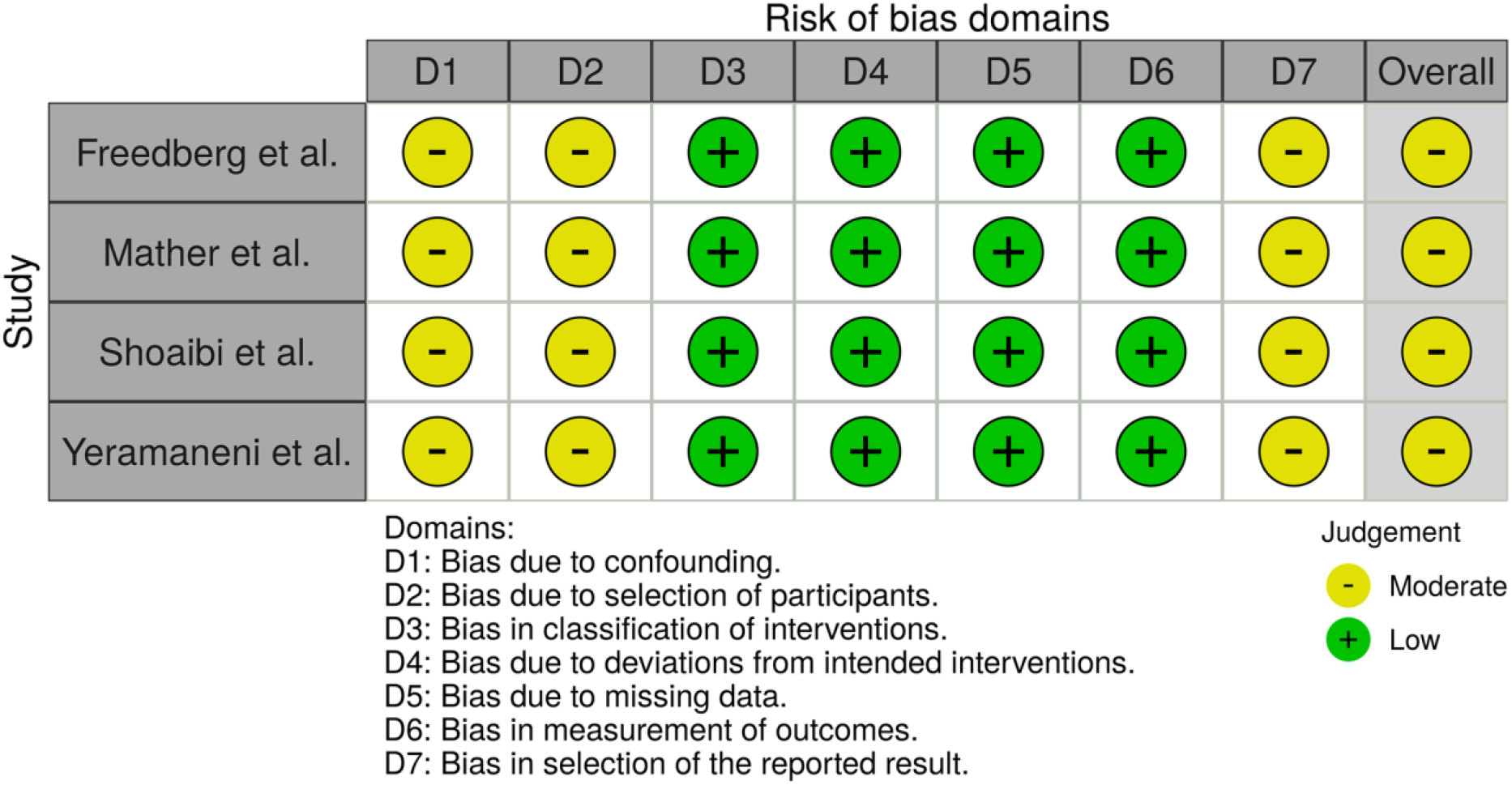
Risk of Bias Assessment

### Clinical Outcomes

Overall, this systematic review and meta-analysis included 46,435 total patients, of whom 3,110 were treated with famotidine during their hospitalizations. Three out of four studies reported a composite endpoint, defined differently in each study but typically consisting of a combination of mortality, intubation, or intensive services (Table 1). Three of four studies reported on rates of mortality, separate from other endpoints.

Across the three studies, COVID-19 patients who took famotidine during hospitalization had a risk of composite outcome that was not significantly different from non-famotidine users by aggregate HR 0.63 (95% CI: 0.35, 1.16; I^2^ = 83.69%). Mather *et al*, the only study that also reported an OR, found a decreased risk of composite outcome: OR 0.47 (95% CI: 0.23, 0.97) (Figure 2a). For Mather *et al*, COVID-19 patients treated with famotidine prior to their hospitalizations were also included; they reported that famotidine use was associated with a lower risk for a composite outcome - HR 0.50 (95% CI: 0.31, 0.79) and OR 0.47 (95% CI: 0.23, 0.97) (Figure 2a) (15).

**Figure 2.**
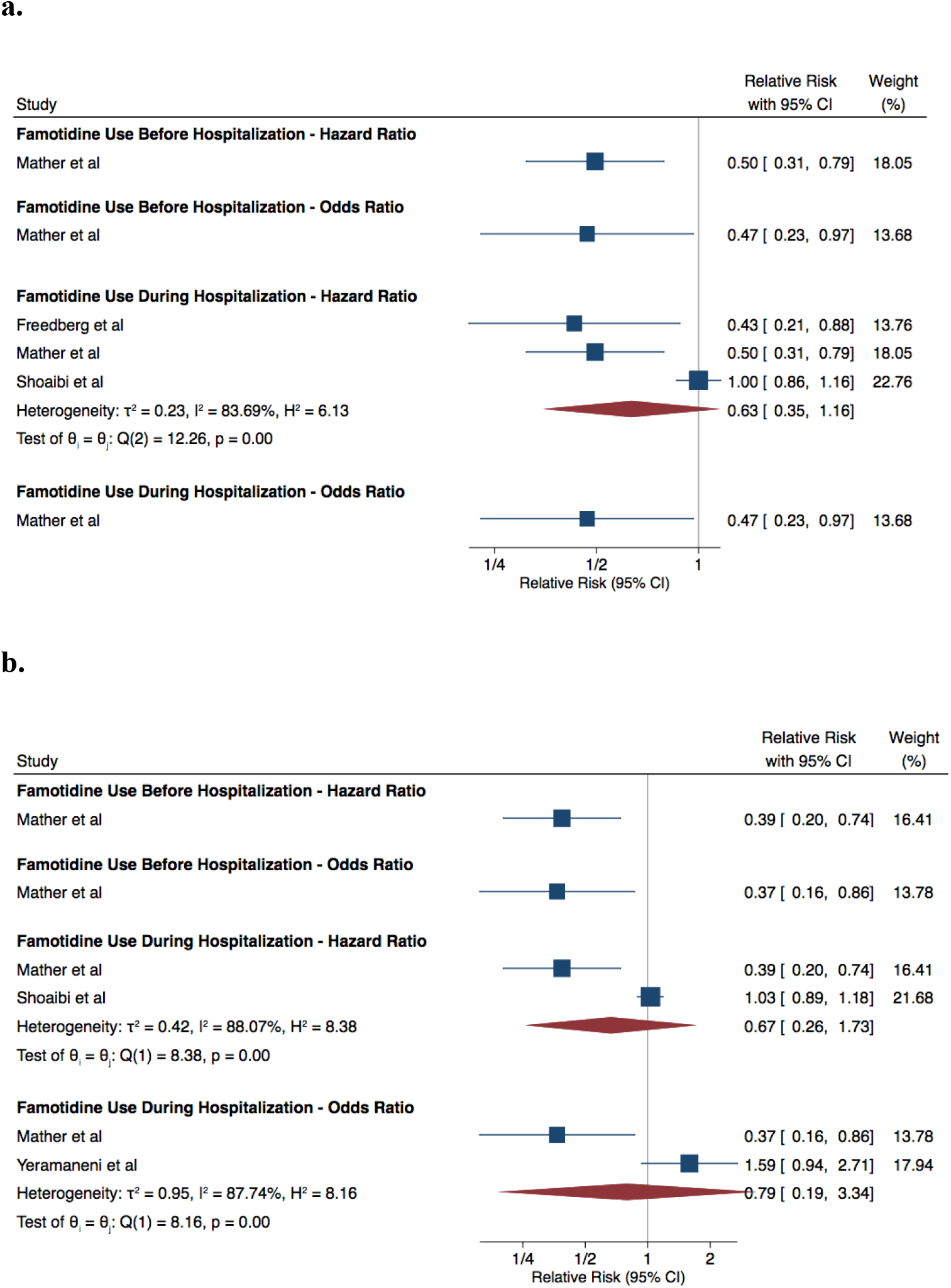
Relative Risk, Compared to Standard of Care **2a**. Composite Outcomes **2b**. Mortality Outcome

Across the three studies that reported mortality rate separated from other endpoints, patients with COVID-19 who received famotidine during hospitalization had mortality rates that did not significantly differ from those who did not receive famotidine - HR 0.67 (95% CI: 0.26, 1.73; I^2^ = 88.07%) and OR of 0.79 (95% CI: 0.19, 3.34; I^2^ = 87.74%) (Figure 2b). Again, Mather *et al*, which included patients who used famotidine before or during COVID-19 hospitalization, reported a decreased risk of mortality - HR of 0.39 (95% CI: 0.20, 0.74) and OR of 0.37 (95% CI: 0.16, 0.86) (Figure 2b). Overall, there was considerable heterogeneity across studies as evidenced by the high I^2^ that ranged from 83.69% to 88.07% depending on analysis.

## DISCUSSION

We conducted an updated systematic review and meta-analysis on the therapeutic impact of famotidine in treating hospitalized COVID-19 patients in accordance with methodological standards set by the MOOSE group for high-quality systematic review and meta-analysis (22). Our results from analyzing four retrospective cohort studies suggest that there is no association between famotidine use and risks of composite outcome of mortality, intubation, and/or use of intensive services, or mortality alone in hospitalized patients with COVID-19. However, the point estimate suggests a direction towards an association with decreased risk of the composite outcome among famotidine users.

Our current analysis is different from the two meta-analyses published by Kamal *et al* and Sun *et al* for four main reasons: 1) to reduce discrepancies in patient characteristics, we did not include the studies by Cheung *et al* and Zhou *et al* as they included all patients with COVID-19 in Hong Kong, including outpatient, inpatient, and emergency settings as opposed to four other studies where only hospitalized patients were included (19, 20); 2) moreover, since both studies drew from the same centralized Hong Kong database, some patients would be counted twice within the same time period if both were included in a meta-analysis as in Kamal *et el* (19); 3) additionally, we chose not to meta-analyze the results of studies that presented a composite endpoint with the results of studies that presented only an endpoint of mortality since they represent different degrees of severity (19, 20); 4) finally, we did not meta-analyze OR along with HR reported by different studies since the rare event assumption is not met (20). In fact, across four studies, the incidence of the composite outcome ranged from 21% to 37% (14-17).

Among reports included in this meta-analysis, two earlier retrospective cohort studies reported that famotidine is associated with a decreased rate of mortality and/or composite outcome for patients with COVID-19 (14-15). For instance, Freedberg *et al* showed a HR of 0.43 (95% CI: 0.21 − 0.88) for the composite outcome of 30-day mortality or intubation when patients were given famotidine on day 1 of hospitalization (14). However, these two studies were single-center studies, had a relatively small sample size of treatment group, and there is a lack of adjustment for concurrent medication use such as corticosteroids, hydroxychloroquine, and azithromycin. On the other hand, two recent studies with larger sample size reported a lack of reduction in mortality and/or composite outcome for famotidine users with COVID-19 (16-17).

Although we did not include Zhou *et al* in this meta-analysis as the study included patients diagnosed with COVID-19 in the ambulatory and emergency settings, it is important to note that the study showed an increased composite outcome of ICU admission, intubation, and all-cause mortality (HR: 1.84, 95% CI: 1.16 −2.92) for COVID-19 patients treated with famotidine (18). The study by Zhou *et al* also investigated the effect of another class of acid suppressor agents in proton pump inhibitors (PPIs), which block the hydrogen/potassium adenosine triphosphatase enzyme system as opposed to the H2 receptors in famotidine. They found that current or regular PPI users were more likely to have severe outcomes of COVID-19 compared to non-users (18) — findings that confirm the results of a meta-analysis in the use of PPIs in patients with COVID-19 (23). The mechanism for this increased risk remains unclear; preliminary hypotheses include PPIs may reduce the secretion of gastric acid that can neutralize the SARS-CoV-2. Despite the use of both PPIs and H2 blockers in the setting of acid suppression, H2 blockers like famotidine may have a better long-term safety profile; observational studies such as one by Yan Xie *et al*. showed that when compared to patients on H2 blockers, patients on PPIs have an increased risk of death (24). Similarly, among the four studies included in this meta-analysis, none showed an increased risk of severe outcomes among COVID-19 patients treated with famotidine. Nevertheless, there remains an urgent need for randomized controlled trials (RCTs) to elucidate the treatment effect and safety profile of famotidine in hospitalized patients with COVID-19. Fortunately, recruitment for a multicentered RCT has been completed and we await its results (25).

Currently, there is very limited data on the efficacy of oral famotidine in treatment of COVID-19 patients with mild to moderate disease solely in the outpatient setting. A case series of 10 non-hospitalized COVID patients reported improved symptoms score after initiation of high dose famotidine (26). However, a survey study conducted in otolaryngology patients found that chronic famotidine use was not associated with incidence of COVID-19 (27). Regardless, higher quality studies such as RCTs are needed to further elucidate the role of famotidine in treating mild to moderate, non-hospitalized COVID-19 patients, and one such study is currently underway at Northwell Health (28).

There are several limitations to this study. First, the strength of our findings is limited by the quality of included studies as is the case for all systematic reviews and meta-analyses. To account for confounding, this meta-analysis contains only observational data that reported adjusted relative risks. Although all the included observational studies had some concern for risk of bias, they employed propensity score matching to minimize selection bias. Additionally, only one study explicitly included patients with COVID-19 treated with famotidine before and during hospitalization (15). Other studies may have included patients who also used famotidine before hospitalization as they may have used as continuation of home use—an assumption made by Freedberg *et al*. Furthermore, while we employed a random effects model for our analysis, the heterogeneity is high. Lastly, there are subtle yet meaningful differences in the definition of composite outcome across the four studies, thereby allowing for potential bias when calculating aggregate ORs/HRs. Given the paucity of data reported in the literature, the directionality of these results should only be used for hypothesis-generation rather than clinical decision making.

In conclusion, this meta-analysis suggests that famotidine does not reduce the risk of mortality in individuals hospitalized with COVID-19. Similarly, there was a point estimate suggesting a decreased risk of the composite outcome of death, intubation, and/or use of intensive services among famotidine users, but this did not meet statistical significance. Further RCTs may help determine the efficacy and safety of famotidine in treating COVID-19 patients in various care settings.

## Data Availability

N/A

## Appendix 1. Search Strategy

Database: Ovid MEDLINE(R) ALL <1946 to February 10, 2021>

1. (covid 19 or covid-19).mp. (98012)
2. “coronavirus disease 2019”.mp. (18967)
3. SARS-CoV-2.mp. (39939)
4. severe acute respiratory syndrome coronavirus 2.mp. (39862)
5. 1 or 2 or 3 or 4 (101636)
6. exp Famotidine/ (1613)
7. “famotidine”.mp. (2318)
8. 6 or 7 (2318)
9. 9 5 and 8 (33)

Database: Embase <1974 to 2021 February 11>

1. (covid 19 or covid-19).mp. (87363)
2. “coronavirus disease 2019”.mp. (87063)
3. SARS-CoV-2.mp. (31402)
4. severe acute respiratory syndrome coronavirus 2.mp. (27219)
5. 1 or 2 or 3 or 4 (101544)
6. exp Famotidine/ (9183)
7. “famotidine”.mp. (9369)
8. 6 or 7 (9369)
9. 9 5 and 8 (66)

Database: EBM Reviews - Cochrane Central Register of Controlled Trials <January 2021>

1. (covid 19 or covid-19).mp. (3882)
2. “coronavirus disease 2019”.mp. (1027)
3. SARS-CoV-2.mp. (213)
4. severe acute respiratory syndrome coronavirus 2.mp. (645)
5. 1 or 2 or 3 or 4 (3988)
6. exp Famotidine/ (464)
7. “famotidine”.mp. (972)
8. 6 or 7 (972)
9. 5 and 8 (7)

Database: medRxiv <February 12, 2021>

1. (covid 19 OR coronavirus 19) AND famotidine (3)

Database: researchsquare <February 12, 2021>

1. (covid 19 OR coronavirus 19) AND famotidine (0)

## Appendix 2. PRISMA Flow Diagram

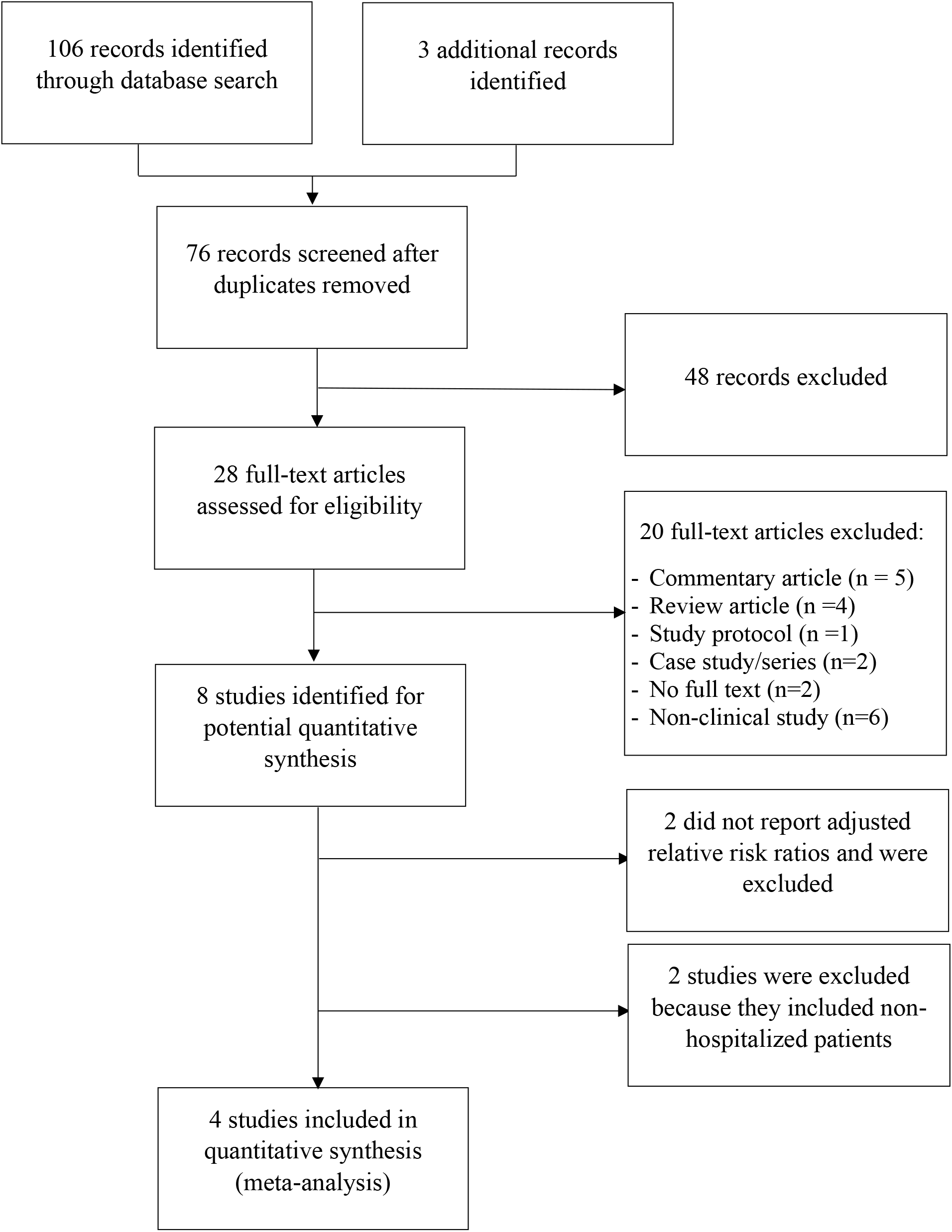

